# Nocturnal cough as a syndromic surveillance signal for respiratory illness in England

**DOI:** 10.64898/2026.07.20.26357937

**Authors:** Tommy Irons, Emil Carlsson, Maria Tang, Jonathon Mellor, Conrad Rubin, Alex Allen, Alex J. Elliot, Mikael Kågebäck, Josef Packham

## Abstract

We evaluated aggregated, privacy-preserving smartphone-detected nocturnal cough activity from the Sleep Cycle application as a potential syndromic surveillance signal in England. Weekly cough metrics from January 2023 to January 2026 were compared with UK Health Security Agency indicators: NHS 111 acute respiratory infection (ARI) triage calls, influenza and COVID-19 PCR positivity, and hospital admission rates for influenza, COVID-19, and respiratory syncytial virus. We evaluated total cough counts alongside two population-normalised metrics, coughs per user and coughs per hour of sleep, and assessed temporal relationships nationally and regionally using cross-correlation with prewhitening. The strongest and most consistent associations were observed for NHS 111 ARI triage calls, where population-normalised cough metrics showed raw national correlations of approximately 0.95 and retained prewhitened correlations above 0.55 at lag 0. This indicates that nocturnal cough activity closely tracks short-term variation in an established syndromic surveillance indicator, beyond shared seasonality, long-term trends, and autocorrelation. Similar near-contemporaneous patterns were observed across regions. Population-normalised cough metrics also showed epidemiologically plausible leading associations with pathogen-specific indicators: coughs per hour of sleep peaked one week before influenza PCR positivity, while both coughs per user and coughs per hour of sleep peaked one week before COVID-19 PCR positivity. Hospital-based indicators showed weaker and more heterogeneous relationships, but the normalised cough metrics still showed plausible temporal alignment with influenza and COVID-19 admissions, including contemporaneous associations with influenza admissions and short leading associations with COVID-19 admissions. In contrast, unnormalised total cough counts produced less stable and often non-interpretable lag structures, consistent with sensitivity to variation in observation volume. These findings suggest that passive, near-real-time nocturnal cough monitoring can provide a population-level signal of respiratory symptom burden, with greatest utility as a broad syndromic indicator that complements surveillance sources affected by healthcare-seeking behaviour, laboratory turnaround times, backfilling, and reporting delays.

## Introduction

Respiratory infections remain a major public health challenge, requiring robust and timely surveillance systems to detect emerging trends and support effective intervention. In the United Kingdom, the UK Health Security Agency (UKHSA) undertakes national surveillance of infectious diseases and publishes routine epidemiological reports that incorporate laboratory diagnoses, hospital admissions, community surveillance and syndromic indicators [1]. These systems collectively support the early detection of seasonal respiratory viruses and novel threats.

The growing availability of digital health data presents an opportunity to strengthen and expand these surveillance capabilities. Everyday interactions with smartphones, wearable devices, and digital health applications generate large volumes of passively collected behavioural and physiological data that may provide timely insights into population health dynamics. This digital footprint has given rise to the research field of *digital epidemiology* [2] and researchers have leveraged alternative data sources such as search trends [3], social media [4], participatory symptom reporting [5], and mobility patterns [6] to perform public health surveillance. While many of these signals provide indirect evidence of disease activity through changes in behaviour, emerging digital health technologies also enable direct measurement of physiological symptoms at population scale.

Among candidate digital epidemiology signals, passively detected nocturnal cough is unusual in that it provides a direct measure of respiratory symptom activity that can be collected continuously, passively, and at population scale. Despite these characteristics, its potential utility for public health surveillance remains largely unexplored. One technology that enables such measurement is sleep monitoring applications, which are widely used smartphone-based tools that analyse audio and movement patterns during sleep. Some applications employ machine learning-based audio event detection to identify and quantify cough events, generating high-frequency measurements of nocturnal cough activity without requiring active user participation [7]. Since coughing is a common symptom of many respiratory infections, aggregated nocturnal cough activity may reflect changes in respiratory disease burden occurring within the community.

In this study, we evaluate the temporal relationship between aggregated nocturnal cough data from the Sleep Cycle application and established UKHSA respiratory surveillance indicators in England. Using retrospective datasets from January 2023 to January 2026, we examine how app-detected cough activity relates to NHS 111 acute respiratory infection triage calls, influenza and COVID-19 PCR positivity, and hospital admission rates for COVID-19, influenza, and respiratory syncytial virus (RSV). We compare multiple population-level cough metrics, including total coughs, coughs per user, and coughs per hour of sleep, at national and regional scales, and use cross-correlation and prewhitening methods to assess the strength, timing, and consistency of associations with established surveillance indicators. By examining these relationships, we aim to determine whether nocturnal cough data provide a timely, symptom-based signal of respiratory illness burden, and whether this signal aligns most strongly with broad syndromic indicators rather than pathogen-specific or severe disease outcomes. These findings can inform future prospective evaluation of consumer-generated respiratory health data as a complementary component of public health surveillance.

### Related Work

Digital epidemiology has increasingly explored the use of non-traditional data sources to complement established public health surveillance systems. Among the most successful examples are participatory surveillance platforms, in which individuals voluntarily report symptoms through online or mobile-based systems. Influenzanet [5], established across multiple European countries, demonstrated that self-reported symptom data can provide reliable estimates of influenza-like illness activity that closely track traditional surveillance indicators [8]. Similar approaches have subsequently been adopted elsewhere, including Flu Near You in the United States [9] and the Dutch Infectieradar platform [10], which has been used to estimate acute respiratory infection incidence and improve attribution of respiratory disease burden to influenza and COVID-19 [11]. During the COVID-19 pandemic, app-based symptom surveillance systems such as the COVID Symptom Study further demonstrated the potential of large-scale citizen-generated health data for monitoring disease activity in near real time [12]. Collectively, these studies demonstrate that digital epidemiology signals can provide useful complements to traditional surveillance systems, while also highlighting the importance of population engagement and sustained participation.

A central question when evaluating novel surveillance signals is whether they provide information earlier than established public health surveillance systems. A range of statistical approaches have been used to assess such leading indicator relationships, with cross-correlation analysis being among the most widely adopted. For example, Hripcsak et al. [13] demonstrated that cross-correlation could be used to validate electronic health record-derived influenza surveillance signals against laboratory-confirmed influenza positivity and emergency department activity. In an English context, Mellor et al. [14] evaluated several digital surveillance sources, including Google Trends, NHS 111 triage calls, and the ZOE symptom study, as leading indicators of COVID-19 hospital admissions using cross-correlation, Granger causality, and dynamic time warping. Google Trends and NHS 111 triage calls consistently preceded admissions by 5–20 days across most geographies, although the strength and timing of these relationships varied across spatial scales. Similarly, Zhang et al. [15] demonstrated that Australian influenza surveillance data led influenza activity in the United States, United Kingdom, and China by more than 20 weeks, while local internet search data provided shorter lead times but improved short-term forecasting performance when combined with surveillance data. More recently, Mellor et al. [16] outlined methodological considerations for evaluating leading indicators in epidemic surveillance, emphasising the importance of causal plausibility, temporal aggregation, reporting delays, spatial heterogeneity, and appropriate validation strategies.

More broadly, several studies have argued that no single surveillance system can fully capture the dynamics of respiratory disease activity. Comparing nine surveillance systems for monitoring COVID-19 in England, Brainard et al. [17] found that different systems provided complementary information and varied substantially in their timeliness, representativeness, and sensitivity to changes in disease activity. These findings highlight the value of integrating diverse surveillance signals rather than relying on any individual source in isolation. However, identifying useful complementary signals remains challenging. Bloom et al. [18] noted that complex temporal processes, reporting delays, and autocorrelation structures can generate misleading associations between surveillance indicators, underscoring the importance of rigorous statistical evaluation when assessing potential leading indicators. Similar concerns have been raised in digital epidemiology, where apparently strong associations can arise from changes in user behaviour, data generation processes, and other sources of bias rather than underlying disease activity [19]. Together, these studies motivate the search for novel surveillance signals that provide information not already captured by existing systems while emphasising the need for careful methodological assessment of their utility.

Beyond actively reported symptoms and behavioural indicators such as search activity, social media content, and mobility patterns, increasing attention has been given to passively collected health signals derived from consumer devices. Recent studies have shown that data generated through routine use of smartphones and wearables can provide population-level insights into health and disease dynamics. For example, Radin et al. [20] demonstrated that aggregated Fitbit-derived resting heart rate and sleep duration improved real-time estimation of influenza-like illness activity in the United States. During the COVID-19 pandemic, several studies further showed that wearable-derived physiological and behavioural signals could detect infection-related changes before or near symptom onset. Mishra et al. [21] found that smartwatch data could identify COVID-19-related physiological changes prior to symptom onset in many individuals, while Quer et al. [22] demonstrated that combining wearable sensor measurements with self-reported symptoms improved COVID-19 detection. However, most existing work has focused on wearable-derived measures such as heart rate, physical activity, or temperature rather than directly observed respiratory symptoms. Nocturnal cough activity therefore represents a related but distinct class of consumer-generated health signal, offering a direct measure of respiratory symptom burden that can be collected passively and continuously at population scale.

The nocturnal cough dataset considered in this study has previously been used to investigate environmental and respiratory health outcomes. Mascolo et al. [23] demonstrated associations between air pollution exposure and nocturnal coughing using aggregated Sleep Cycle cough data. More broadly, recent work on cough monitoring has highlighted the potential of passively measured cough activity as a marker of respiratory health and disease burden [7]. While these findings suggest that passively collected nocturnal cough activity may provide a useful population-level respiratory health signal, little is known about how such signals relate to established respiratory surveillance indicators. In particular, relationships with pathogen-specific indicators, syndromic surveillance measures, and severe disease outcomes have not been characterised. The present study addresses this gap using respiratory surveillance data from England.

## Results

We evaluated weekly associations between nocturnal cough metrics and established respiratory surveillance indicators at national and regional levels (see Figure 1 for an illustrative example). For each outcome, we compared total cough counts with two population-normalised metrics: coughs per user and coughs per hour of sleep. Cross-correlations were examined before and after prewhitening, with the prewhitened results used as the primary basis for interpretation. Prewhitening removes temporal structure from the cough series before evaluating cross-correlations, reducing the risk that apparent relationships arise solely from shared seasonality, long-term trends, or autocorrelation rather than a genuine temporal association between surveillance signals. ARIMA model specifications and prewhitening diagnostics are provided in the supplementary material.

**Fig 1.**
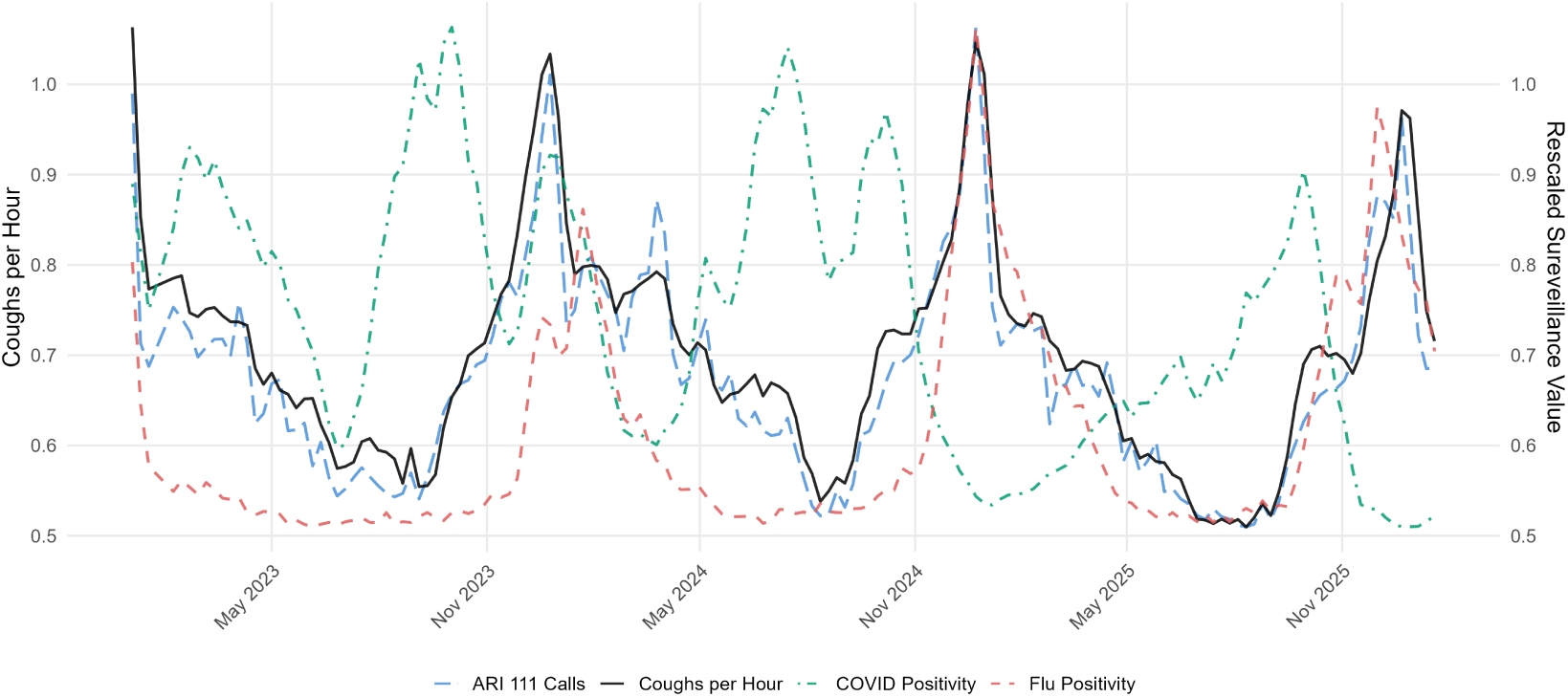
National weekly nocturnal cough activity and selected respiratory surveillance indicators in England, January 2023 to January 2026. Several major peaks in cough activity occur during periods of elevated respiratory disease activity, suggesting that passively collected nocturnal cough data may contain information relevant to public health surveillance. The cough rate (average coughs per hour of sleep) is shown on the left axis; influenza positivity, COVID-19 positivity, and NHS 111 acute respiratory infection (ARI) triage calls are rescaled for visual comparison and shown on the right axis.

Table 1 summarises the national weekly cross-correlation results. The strongest and most consistent relationships were observed for NHS 111 acute respiratory infection (ARI) triage calls. Population-normalised cough metrics showed very high raw correlations with ARI calls and retained substantial prewhitened correlations after removal of temporal structure, with coughs per user and coughs per hour of sleep both peaking at lag 0. This indicates that nocturnal cough activity closely tracked short-term variation in an established syndromic surveillance indicator, beyond shared seasonality and autocorrelation. Similar near-contemporaneous patterns were observed across NHS England regions.

**Table 1.**
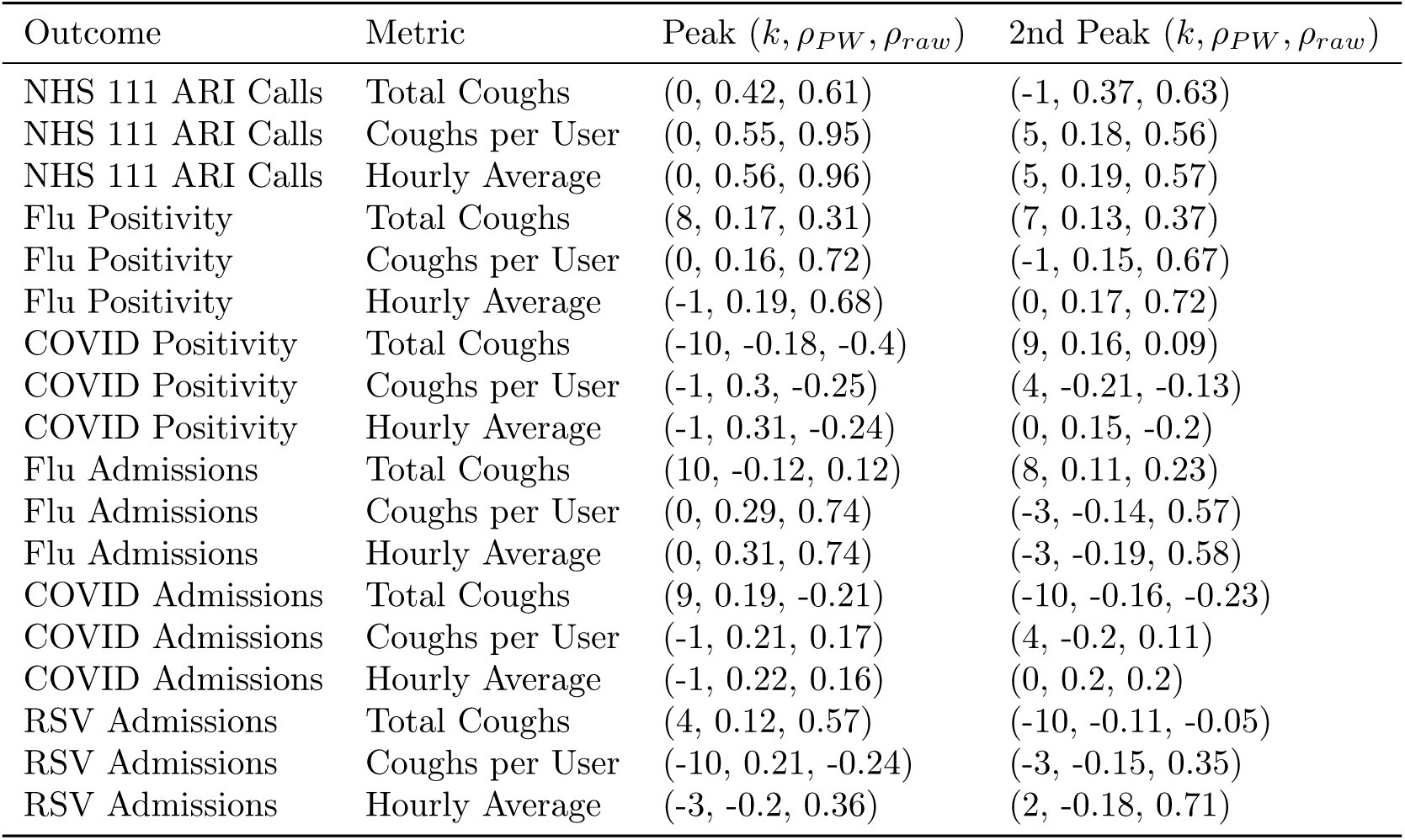
Peak and secondary cross-correlations for national weekly cough series. Values are shown as (lag, prewhitened correlation, raw correlation). Negative lags indicate that cough activity leads the outcome.

Associations with pathogen-specific indicators were more modest but still informative for the population-normalised cough metrics. Influenza PCR positivity showed weak short-term associations, with coughs per user peaking contemporaneously and coughs per hour of sleep peaking at a one-week lead. COVID-19 PCR positivity showed a clearer short leading pattern, with both population-normalised metrics peaking one week before positivity. Hospital admission indicators showed weaker and more heterogeneous relationships, although normalised cough metrics showed contemporaneous associations with influenza admissions and short leading associations with COVID-19 admissions. RSV results were weak and inconsistent, likely reflecting both the shorter available data window and differences between the Sleep Cycle user population and the groups most affected by severe RSV disease.

Across outcomes, the population-normalised metrics more consistently tracked or preceded comparator surveillance indicators, whereas total cough counts produced less stable and less interpretable lag structures, including several peaks at longer or less epidemiologically plausible lags. This likely reflects the sensitivity of raw cough counts to variation in observation volume, including changes in the number of active users and the amount of recorded sleep, rather than changes in respiratory symptom burden alone. Results for total cough counts are therefore presented for completeness, but interpretation focuses primarily on coughs per user and coughs per hour of sleep.

### NHS 111 ARI Triage Calls

Figure 2 presents the cross-correlation analysis between nocturnal cough activity and NHS 111 ARI triage calls. At the national level, coughs per user produced the strongest association observed in the study, with the prewhitened cross-correlation peaking at a lag-0 (*ρ* = 0.55). Total coughs and coughs per hour of sleep showed similar patterns, also peaking at a lag of 0 weeks (*ρ* = 0.42 and *ρ* = 0.56, respectively). Despite the removal of shared temporal structure through prewhitening, the relationship remained clearly visible across all cough metrics, indicating that the association is not solely explained by common seasonality or long-term temporal trends. An approximately Normal 95% confidence interval is shown with upper and lower limits as dotted lines in Figure 2, representing the null hypothesis position of zero correlation. Statistically significant correlations at lags between-1 and 1 are seen across spatial and temporal scales, showing evidence of a relationship between the two series at these lags beyond random variation alone. The regional analyses closely mirrored the national findings. Across the seven NHS England regions, peak correlations consistently occurred at or near lag 0, with relatively little variation in either lag or correlation magnitude.

**Fig 2.**
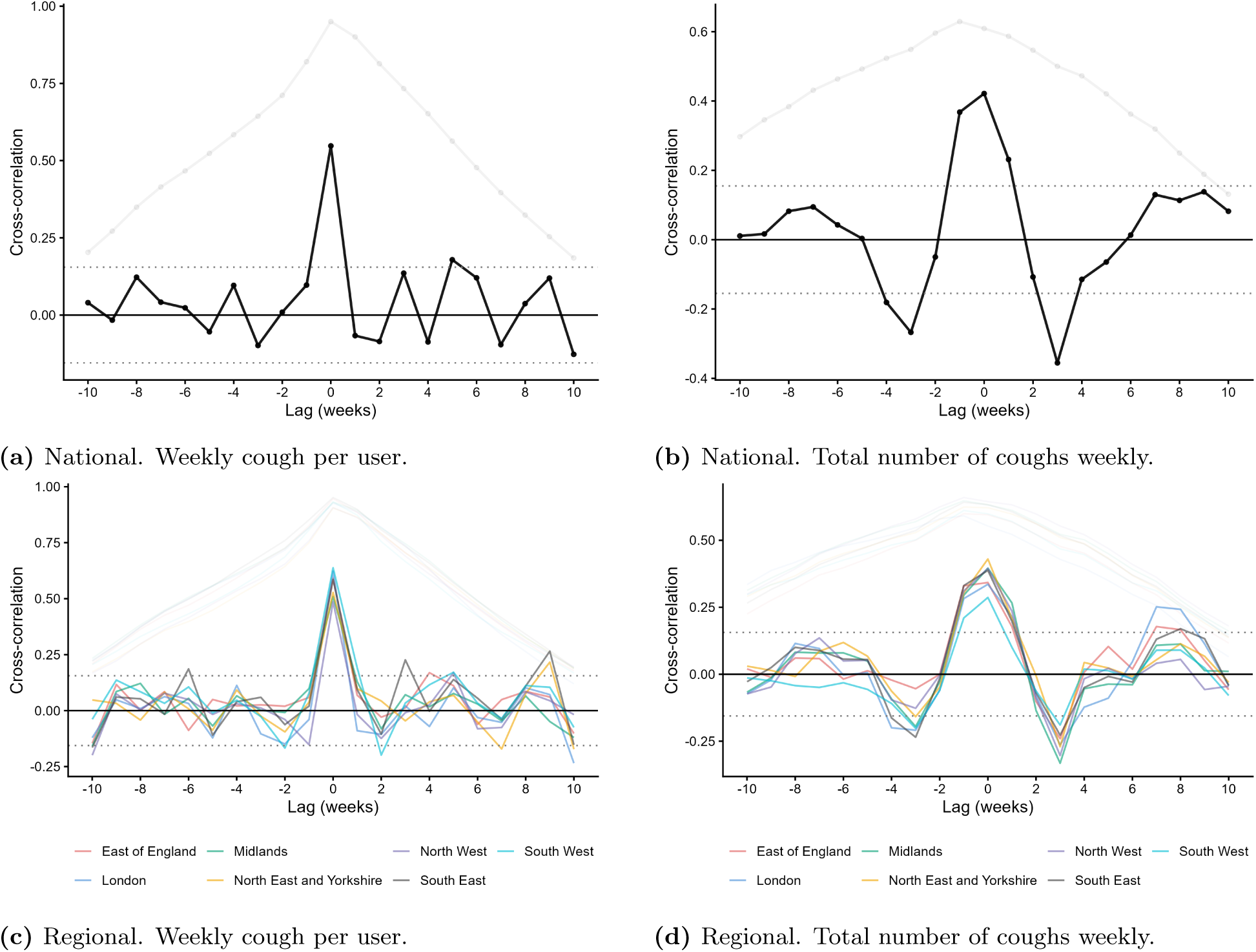
Prewhitened cross-correlation analysis between nocturnal cough activity and NHS 111 acute respiratory infection (ARI) triage calls at national and regional scales. Negative lags indicate that cough activity precedes ARI triage calls. Top row shows result for the metric cough per user during a week while the bottom row shows the results for total number of coughs registered within a week. Normal 95% confidence intervals for the correlations are shown as dotted horizontal lines.

### Influenza

Figure 3 presents the cross-correlation analysis between nocturnal cough activity and influenza PCR positivity. At the national level, the population-normalised cough metrics showed weak but epidemiologically plausible short-term associations with influenza PCR positivity. Coughs per user peaked contemporaneously at lag 0 (*ρ* = 0.16), while coughs per hour of sleep peaked at a one-week lead (*ρ* = 0.19). In contrast, total cough counts peaked at a longer positive lag of +8 weeks (*ρ* = 0.17), a timing that is less epidemiologically interpretable and likely reflects sensitivity to observation volume or longer-term temporal structure in the unnormalised series. Age-stratified analyses showed some statistically significant correlations for the coughs per user metric, but these were not consistent across age groups, with smaller and less clearly significant peaks observed in several strata.

**Fig 3.**
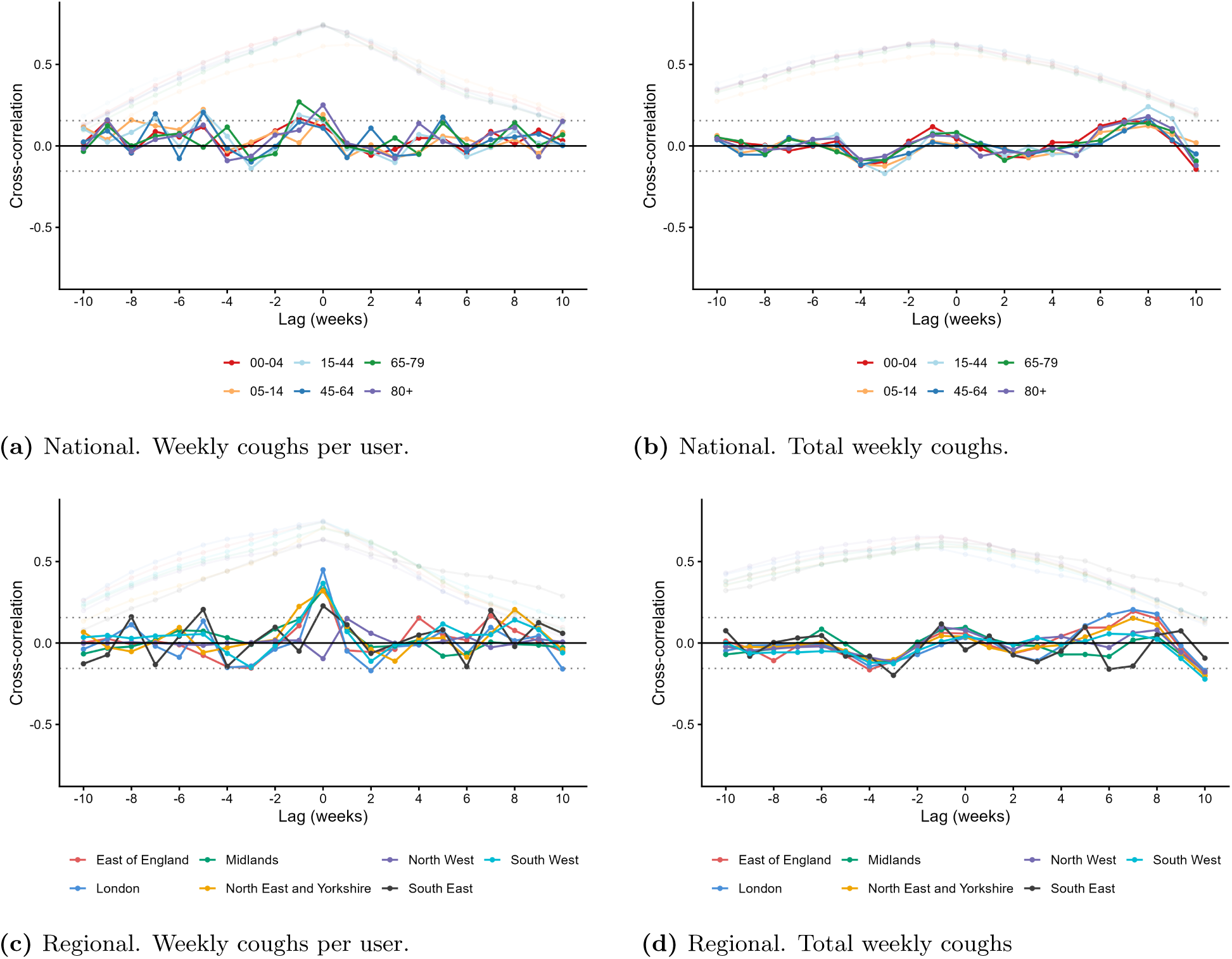
Cross-correlation analysis of weekly cough metrics against influenza PCR positivity, nationally and regionally. Faded lines show raw series; solid lines show prewhitened series. Age groups (national) and NHS regions (regional) are shown as separate coloured lines. Normal 95% confidence intervals for the correlations are shown as dotted horizontal lines.

Regional analyses showed variability in both the magnitude and timing of peak correlations, with different regions exhibiting peaks at negative, zero, and positive lags. However, this variability was less pronounced for the population-normalised metrics, where regional peaks were more often concentrated at or near lag 0. This suggests that the scaled cough metrics captured a weak but temporally plausible influenza-related signal, whereas the lag structure for total cough counts was less stable across regions.

Overall, these results indicate that influenza positivity showed only modest associations with nocturnal cough activity, but that interpretation depended strongly on the cough metric. The population-normalised metrics showed epidemiologically plausible short-term associations, with coughs per user peaking contemporaneously and coughs per hour of sleep peaking at a one-week lead. In contrast, total cough counts peaked at a longer positive lag, suggesting that the unnormalised metric was more sensitive to changes in observation volume or longer-term temporal structure than to influenza activity itself. Thus, while influenza positivity did not show the same strong and consistent relationship observed for NHS 111 ARI triage calls, the normalised cough metrics provide evidence that influenza-related respiratory activity contributes to the broader nocturnal cough signal.

### COVID-19

Figure 4 presents the cross-correlation analysis between nocturnal cough activity and COVID-19 PCR positivity. In contrast to NHS 111 ARI triage calls, the relationship with COVID-19 positivity was more moderate but still showed evidence of a consistent short-term temporal pattern for population-normalised metrics, while total coughs showed inconsistent and non-interpretable lag structure. At the national level, both coughs per user and hourly average exhibited statistically significant peak correlations at a lag of*−*1 week (*ρ* = 0.30 and *ρ* = 0.31, respectively), indicating a modest leading relationship. Total coughs also showed a peak at a negative lag, though at a longer time offset (*−*10 weeks, *ρ* = *−*0.18), suggesting greater sensitivity to longer-term temporal structure and aggregation effects in the unscaled series.

**Fig 4.**
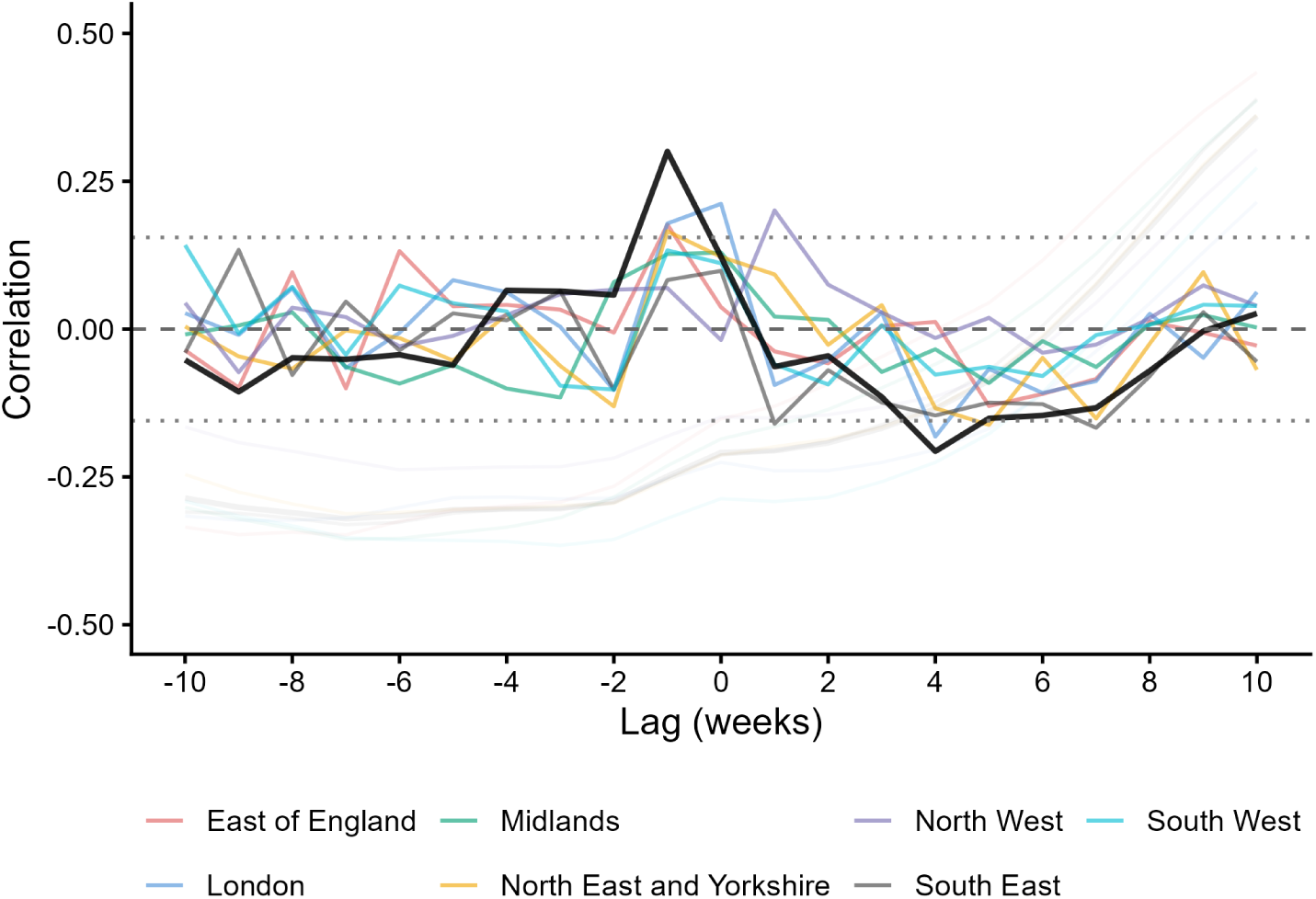
Cross-correlation analysis of weekly coughs per user against COVID-19 PCR positivity. Regional estimates are shown in colour and the national estimate in black. Faded lines show raw series and solid lines show prewhitened series. Negative lags indicate that cough activity precedes COVID-19 positivity. Normal 95% confidence intervals for the correlations are shown as dotted horizontal lines.

Regional analyses showed variability in the exact timing of peak correlations and their associated significances, but peaks for the normalised cough metrics were generally concentrated at negative or near-contemporaneous lags. This suggests that the short-term leading association observed nationally was not driven by a single cough metric, although its magnitude and timing varied across regions. In contrast, the total cough series showed less interpretable behaviour, consistent with greater sensitivity to longer-term temporal structure and variation in observation volume.

Overall, these findings indicate that normalised nocturnal cough metrics showed a weak but coherent short-term leading association with COVID-19 positivity. The relationship was weaker than that observed for NHS 111 ARI triage calls, but it remained positive after prewhitening and was consistent across coughs per user and coughs per hour of sleep at the national level. This suggests that COVID-19-related respiratory activity contributed to the broader nocturnal cough signal, even though nocturnal cough activity is not sufficiently pathogen-specific to serve as a standalone COVID-19 surveillance indicator.

### Hospital Admission Rate

Figure 5 presents the cross-correlation analysis between nocturnal cough activity and hospital admission rates for COVID-19, influenza and RSV. For COVID-19 admissions, a weak short-term leading relationship was observed for the normalised metrics, with both coughs per user and hourly average peaking at a lag of *−*1 week (*ρ* = 0.22 and *ρ* = 0.21, respectively). In contrast, total coughs peaked at a longer positive lag of +9 weeks (*ρ* = 0.19). Regional analyses showed heterogeneity in peak timing, with no consistent pattern of leading behaviour across all regions, although negative and near-contemporaneous lags were common for the scaled metrics.

**Fig 5.**
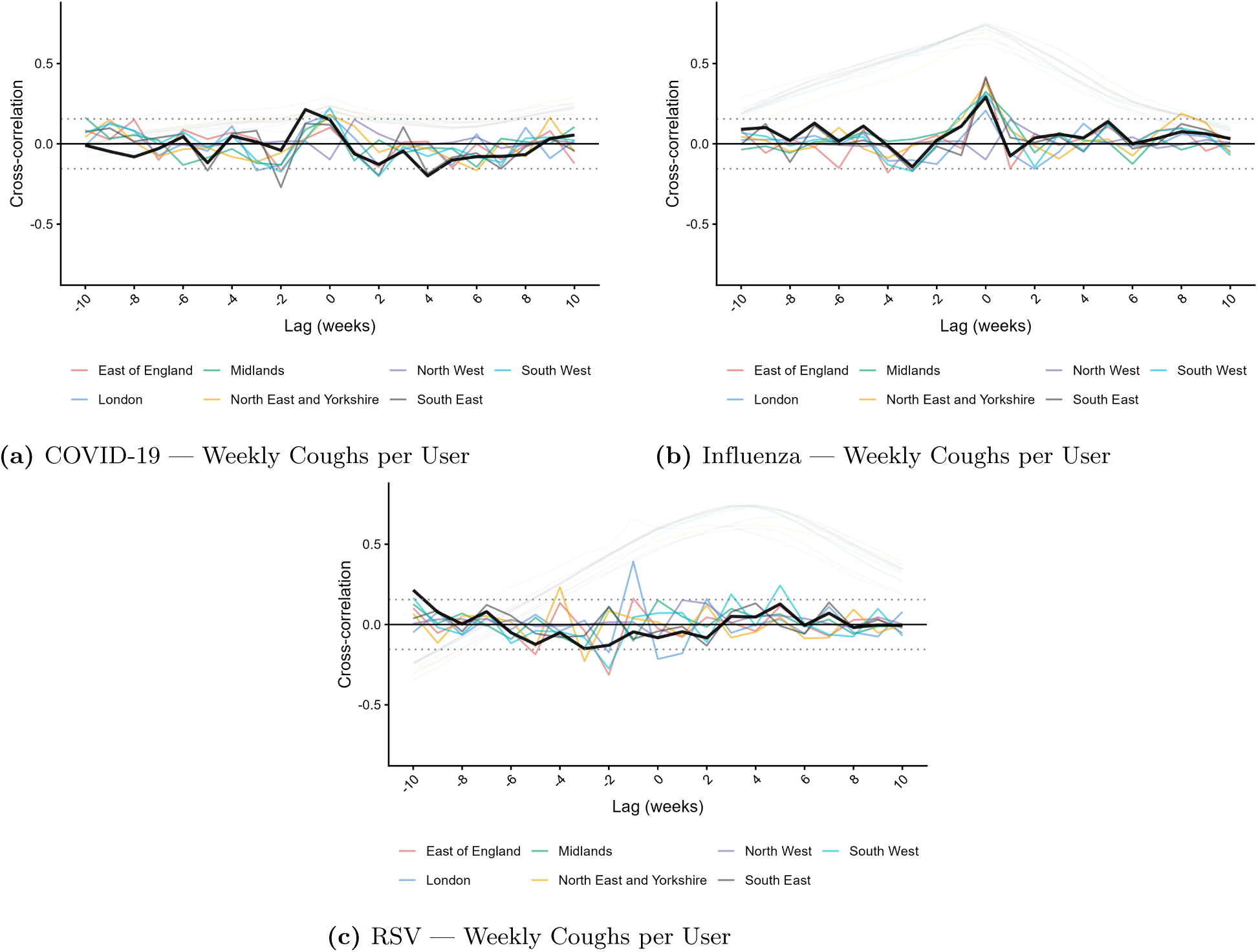
Cross-correlation analysis of coughs per user against hospital admission rates for COVID-19, influenza, and RSV, nationally and regionally. Faded lines show raw series; solid lines show prewhitened series. NHS regions are shown as separate coloured lines with the national estimate overlaid in black. Normal 95% confidence intervals for the correlations are shown as dotted horizontal lines.

For influenza admissions, the normalised cough metrics showed broadly consistent contemporaneous relationships at both national and regional scales. Nationally, coughs per user and hourly average both peaked at a lag of 0 weeks (*ρ* = 0.29 and *ρ* = 0.31, respectively), lying above the confidence intervals and hence are statistically significant findings. Total coughs exhibited a peak at a longer positive lag of +10 weeks (*ρ* = *−*0.12). Regional results showed variability in the timing of peak correlations, with peaks distributed across negative, zero, and positive lags, many of which fall outside reasonable temporal windows and likely reflect residual temporal structure rather than causal relationships.

For RSV admissions, no clear or consistent temporal relationship was observed. Nationally, total coughs showed only a weak peak at a positive lag of +4 weeks (*ρ* = 0.12), while coughs per user peaked at a longer negative lag of *−*10 weeks (*ρ* = 0.21). Coughs per hour of sleep also showed an implausible lag structure, with the largest absolute association occurring at *−*3 weeks but in the negative direction (*ρ* = *−*0.20). Regional results were similarly variable, with no coherent pattern in either lag or correlation magnitude.

To summarize, hospital admission indicators showed weaker and more heterogeneous relationships with nocturnal cough activity than NHS 111 ARI triage calls, as expected for outcomes reflecting more severe disease and additional delays between symptom onset, testing, and admission. However, the normalised cough metrics still showed plausible associations with influenza and COVID-19 admissions: influenza admissions were most strongly associated with contemporaneous cough activity, while COVID-19 admissions showed a weaker short-term leading pattern. In contrast, total cough counts produced less interpretable peak lags, again suggesting sensitivity to observation volume and longer-term temporal structure. RSV admission results were not sufficiently consistent to support interpretation. This likely reflects both the shorter available RSV time series and differences between the Sleep Cycle user population and the groups most affected by severe RSV disease, particularly infants and young children, who are under-represented in the cough dataset (see Figure 6).

**Fig 6.**
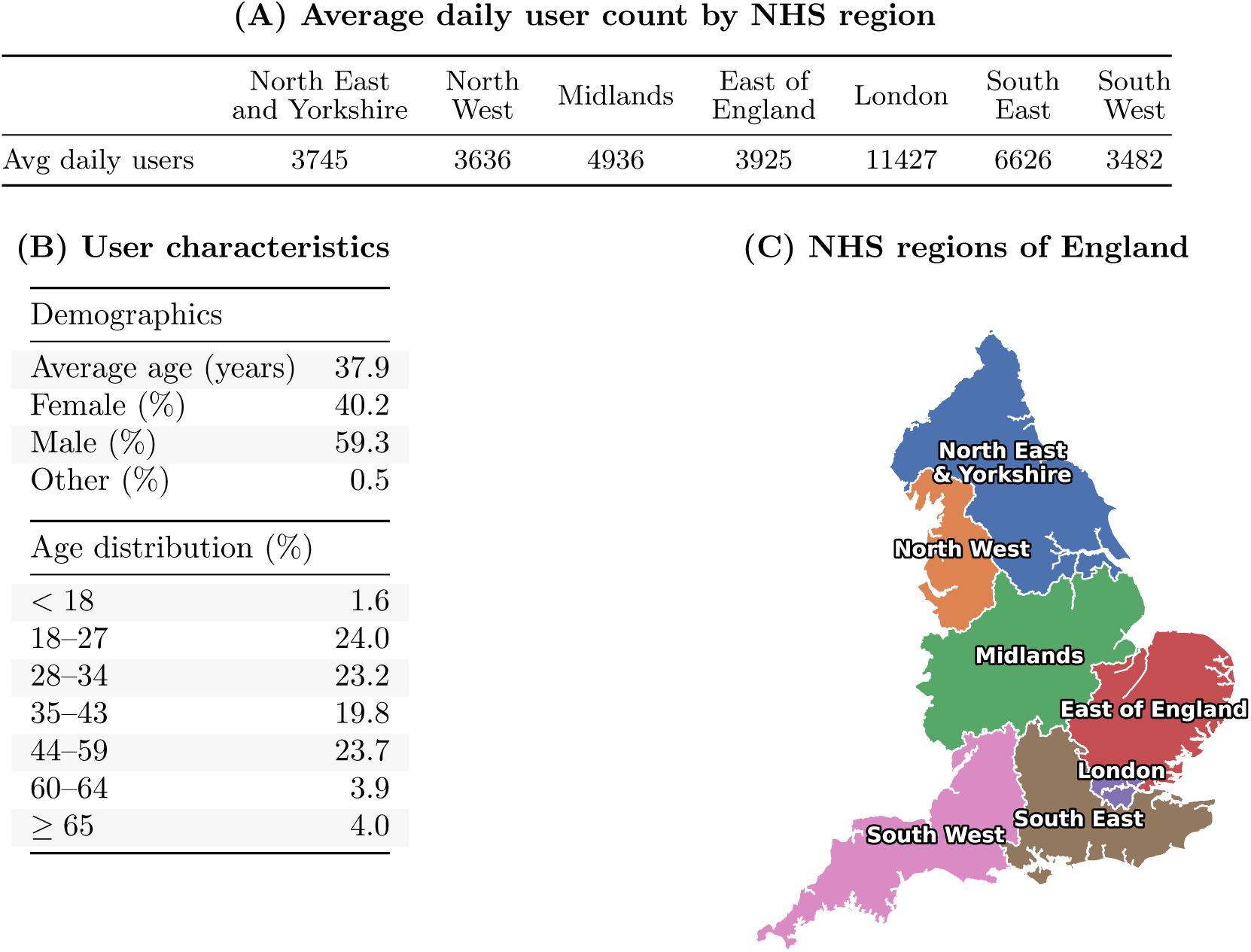
Overview of study population and spatial aggregation. **(A)** Average daily active users reporting to the cough dataset across the seven NHS regions. **(B)** Characteristics of the users in the cough dataset. Reporting these values is optional for users; the characteristics presented here should therefore be interpreted as estimates **(C)** Geographical boundaries of the NHS regions.

## Discussion

### Nocturnal cough as a syndromic surveillance signal

This study evaluated whether passively collected nocturnal cough activity from a consumer sleep monitoring application could provide a timely signal of respiratory disease activity in England. The strongest and most consistent relationships were observed for NHS 111 ARI triage calls, where population-normalised cough metrics closely tracked call volume at both national and regional scales. Importantly, the association with NHS 111 ARI calls persisted after prewhitening, suggesting that the cough signal captures short-term variation in respiratory symptom burden beyond shared seasonal structure. The consistency of the ARI association across NHS England regions suggests potential value for regional situational awareness, despite demographic and geographic skew in app users.

Associations with pathogen-specific indicators were generally more moderate but still informative: COVID-19 positivity and admissions showed weak to moderate short leading associations for normalised cough metrics, influenza admissions showed moderate contemporaneous associations, and influenza positivity was weaker but with some leading and contemporaneous associations. RSV admission results were inconclusive due to the shorter available time series.

These findings suggest that nocturnal cough activity functions primarily as a broad syndromic indicator of respiratory illness burden rather than a pathogen-specific surveillance signal. This pattern is biologically plausible: cough is a common symptom across multiple respiratory infections, but its timing, severity, and persistence vary by pathogen, variant, host factors, and disease severity. As a result, aggregated nocturnal cough activity is unlikely to map cleanly onto any single pathogen-specific indicator. Instead, its value appears greatest as a direct, passively measured signal of community respiratory symptom burden.

The results also highlight the importance of how nocturnal cough activity is represented. Across several outcomes, the population-normalised metrics, coughs per user and coughs per hour of sleep, produced more coherent and interpretable associations than total cough counts, with peak correlations more consistently occurring at lags where cough activity either coincided with (lag 0) or moderately preceded (typically 1 week) the comparator surveillance indicators. This is expected because total cough counts reflect both changes in respiratory symptom burden and changes in observation volume, including variation in the number of active users and the amount of recorded sleep. In contrast, coughs per user adjusts for changes in the active user population, while coughs per hour of sleep also accounts for variation in recorded sleep duration. The less stable lag structure observed for total cough counts, including peaks at longer or less epidemiologically plausible lags for some outcomes, likely reflects this sensitivity to changes in data volume rather than respiratory illness dynamics alone. While total cough counts were included for completeness, their frequent association with long or implausible lag structures indicates that they should not be used as a surveillance signal in practice. For surveillance applications, population-normalised cough metrics therefore appear more suitable than raw cough counts, as they better isolate changes in cough intensity from changes in platform usage or observation time and more consistently align with subsequent or contemporaneous changes in established surveillance indicators.

Moreover, our findings fit within a broader view of respiratory surveillance as an ecosystem of complementary, imperfect indicators rather than a hierarchy of interchangeable signals. Brainard et al. [17], comparing multiple COVID-19 surveillance systems in England, emphasised that no single system could meet all surveillance needs and that different indicators varied in timeliness, representativeness, sensitivity, and susceptibility to behavioural or operational influences. The present findings support a similar interpretation for nocturnal cough monitoring. The strong association with NHS 111 ARI triage calls suggests that passively measured cough captures a meaningful component of the community respiratory symptom burden reflected in an established syndromic surveillance indicator. At the same time, the two signals arise through different data-generating mechanisms: NHS 111 calls reflect symptoms severe or concerning enough to prompt healthcare-seeking, whereas nocturnal cough activity is measured passively during routine app use. This distinction is central to the potential value of cough monitoring. Unlike call-based indicators, the cough signal does not require individuals to recognise symptoms, decide to seek advice, or interact with the healthcare system, and may therefore be less directly affected by changes in public awareness, media attention, service availability, or demographic and socioeconomic differences in healthcare-seeking behaviour. In addition, the near-real-time availability of the cough data, with a reporting lag of less than one day, compares favourably with existing UKHSA sources, which exhibit a normal reporting lag of several days. These characteristics suggest nocturnal cough signals may provide near-real-time nowcasting or rapid corroboration of rises in community respiratory symptoms, and its operational timeliness may still produce practical warning relative to delayed surveillance feeds.

### Limitations

Several limitations should be considered when interpreting these findings. First, the Sleep Cycle user population is not representative of the general population of England. Users tend to be younger and somewhat concentrated in urban areas, particularly London and southern England. Although leading relationships were observed across multiple regions, this demographic and geographic skew may influence the observed associations, e.g., the young age profile of app users may attenuate relationships with hospital admissions, particularly for RSV and influenza, where severe disease burden is concentrated in older adults and young children.

Second, this study evaluates retrospective temporal associations rather than prospective predictive performance. Although cross-correlation and prewhitening provide useful tools for assessing temporal relationships, they do not establish operational utility. Demonstrating practical surveillance value would require prospective evaluation and comparison against forecasting baselines in a real-time setting.

Third, the analyses presented here were conducted using surveillance data as reported and did not explicitly adjust for reporting delays in those signals. Because nocturnal cough activity is available with minimal delay, whereas comparator surveillance systems may incorporate delays arising from healthcare utilisation, laboratory processing, reporting workflows, and data consolidation, the observed temporal patterns may represent conservative estimates of the true temporal relationship. In other words, some surveillance indicators may only become available several days after the underlying health events they measure. While the analyses were conducted at weekly resolution, meaning that (relatively) small reporting delays are unlikely to materially alter the overall conclusions, explicitly accounting for reporting delays would provide a more accurate assessment of the operational early-warning value of nocturnal cough monitoring.

Several methodological considerations should also be acknowledged. Prewhitening substantially reduced the influence of shared temporal structure and autocorrelation, but like all time-series approaches, it relies on modelling assumptions that may not fully capture complex epidemiological dynamics.

Finally, interpretation of the RSV analyses is limited by the shorter available time series, which covered substantially fewer respiratory seasons than the other outcomes. The study was also conducted during a post-pandemic period characterised by changing respiratory virus circulation patterns and healthcare behaviours, which may limit generalisability to future seasons or other countries.

### Future Directions

Future research should explore prospective evaluation of nocturnal cough activity within an operational surveillance setting. While the present study demonstrates temporal relationship with several established surveillance indicators, it remains unclear whether these relationships would translate into actionable early warning in real time. Integrating cough data into existing surveillance dashboards and evaluating performance across future respiratory seasons would provide a useful next step.

Other avenues for research include exploring temporal variation in the cough–surveillance relationship across seasons and pathogens, evaluating performance in other countries and healthcare systems, and investigating whether demographic stratification or more advanced signal processing approaches can improve specificity.

An additional area for future research is the integration of other passively collected respiratory signals available through sleep monitoring applications. These applications can estimate physiological measures including breathing rate during sleep. Changes in respiratory rate are associated with a range of respiratory infections and may capture different aspects of disease progression than cough alone. Exploring combinations of cough activity, breathing rate, sleep disruption, and other passively measured signals may therefore yield more robust and informative surveillance indicators than any individual metric in isolation. Such multimodal approaches could potentially improve sensitivity to emerging respiratory disease activity while providing a richer picture of population respiratory health.

### Concluding Remarks

Passively collected nocturnal cough activity showed strong alignment with NHS 111 ARI calls in England, supporting its interpretation as a timely, symptom-based measure of community respiratory illness burden. Associations with pathogen-specific and severe disease indicators were generally weaker, but nocturnal cough activity still showed plausible leading or contemporaneous relationships with influenza and COVID-19 outcomes. This pattern suggests that pathogen-specific respiratory activity can contribute to the cough signal, while also confirming that aggregated cough activity is not specific to any single infection. Overall, nocturnal cough is best understood as a broad syndromic indicator rather than as a proxy for an individual pathogen. While our results indicate a relationship between nocturnal cough activity, passively measured by a smartphone application, and population-level respiratory illness burden, prospective evaluation is needed to determine whether nocturnal cough monitoring improves operational surveillance utility, including short-term forecasting and nowcasting. Nevertheless, these findings suggest that consumer-generated respiratory health data may provide a valuable complementary source of information for public health surveillance. As smartphone-based health technologies continue to expand, passively collected respiratory signals such as cough activity and breathing rate may become increasingly useful components of future surveillance systems.

## Materials and methods

### Study design and setting

We conducted a retrospective observational study evaluating aggregated nocturnal cough activity as a potential respiratory surveillance signal in England. Three representations of cough activity - total coughs, coughs per user, and coughs per hour of sleep - were compared against established UKHSA surveillance indicators spanning pathogen-specific measures (influenza and COVID-19 PCR positivity), syndromic surveillance (NHS 111 acute respiratory infection triage calls), and severe disease outcomes (hospital admissions for COVID-19, influenza, and respiratory syncytial virus). Analyses were conducted at both national and regional scales across the seven NHS England regions. The study period ran from 2 January 2023 to 25 January 2026, chosen to maximise temporal overlap between the Sleep Cycle and UKHSA datasets. All analyses were conducted using weekly aggregated time series to reduce the influence of day-to-day variability and reporting artefacts commonly present in public health surveillance data.

### Sleep Cycle nocturnal cough data

We used nocturnal cough data collected through the Sleep Cycle smartphone application, a commercially available sleep-tracking application available on major mobile platforms. Sleep Cycle passively analyses audio recorded during user-initiated sleep sessions, enabling the measurement of nocturnal cough activity without requiring active symptom reporting or other user input. For each sleep session, the application records the total number of detected cough events and the duration of recorded sleep. These measurements were subsequently aggregated across users to produce population-level indicators of nocturnal cough activity.

Cough events are identified using a machine learning–based audio event detection model operating locally on the user’s device. During active sleep-tracking sessions, the model performs inference on overlapping 10-second audio clips and identifies cough events throughout the night. Consecutive cough detections separated by less than one second are treated as a single cough event. At the end of each sleep session, the model reports the total number of detected cough events together with the total duration of recorded sleep.

All audio processing is performed locally on the user’s device and no raw audio recordings are transmitted to Sleep Cycle servers. To preserve user privacy, cough records are anonymised prior to transmission through removal of personal identifiers and perturbation of geographic coordinates using independent noise drawn from a zero-centred Laplacian distribution applied separately to latitude and longitude coordinates.

Cough records were spatially aggregated to the seven NHS England regions using anonymised user location data. Figure 6 summarises the regional coverage and demographic characteristics of the Sleep Cycle user population included in the study. The average daily number of users ranged from 3,482 in the South West to 11,427 in London. Among users who had voluntarily provided demographic information, the average age was 37.9 years, 40.2% were female, 59.3% were male, and 0.5% reported another gender.

### Cough metrics

To assess whether different representations of nocturnal cough activity influence its utility as a surveillance signal, we evaluated three complementary cough metrics. Total cough counts capture the overall volume of detected cough activity but may be influenced by variation in the number of active users over time. We therefore additionally considered population-normalised measures based on the number of active users and the total duration of recorded sleep.

Total coughs were defined as the sum of all cough events detected across all users in a given time period:

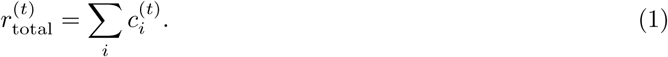

The average hourly cough rate was defined as the ratio of total coughs to the total hours of sleep recorded in the same period:

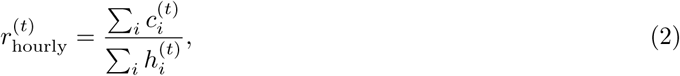

where *c*^(*t*)^ is the number of coughs recorded for user *i* in period *t*, and *h*^(*t*)^ is the corresponding number of hours of sleep. Coughs per user were defined as the ratio of total coughs to the number of active users in the same period:

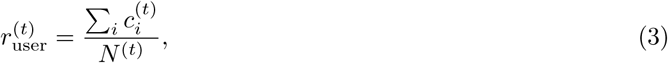

where *N* ^(*t*)^ is the number of active users in period *t*. Each metric was computed at national and regional levels and at weekly temporal resolution.

### Public health surveillance data sources

#### NHS 111 acute respiratory infection triage calls

UKHSA undertakes daily surveillance of NHS 111 triaged call data to provide early warning of health threats or situational awareness during health emergencies [24]. NHS 111 acute respiratory infection triage (ARI) calls were included as a syndromic indicator. NHS 111 is a non-emergency telephone service in England that members of the public can contact when they need medical advice or assistance but the situation is not life-threatening. Callers are triaged by trained advisors using a clinical decision support system, and directed to the most appropriate care. Because 111 captures a high volume of patient-initiated contacts at the point when symptoms first prompt someone to seek help, call data for conditions like ARIs serve as a near-real-time indicator of community illness burden. Daily counts of NHS 111 triage calls categorised as relating to an ARI were obtained from the UKHSA Syndromic Surveillance team. Data are available at both national and regional level. For regional analyses, data provided at a finer geographic resolution were aggregated to the seven NHS England regions. Weekly totals were derived by summing daily counts within each Monday-to-Sunday week.

#### Influenza and COVID-19 PCR positivity

Influenza and COVID-19 positivity were included as pathogen-specific indicators of respiratory virus circulation. Positivity is defined as the proportion of samples testing positive among all submitted for PCR testing for a given pathogen. The daily regional influenza PCR testing positivity data were a product of the second generation surveillance system (SGSS), the UKHSA’s national laboratory reporting system for infectious diseases covering all of England across all clinical settings. No scaling was applied for the regional analysis, while the SGSS data were aggregated over the regions for national evaluations on a daily scale.

Weekly influenza PCR positivity data were obtained from the UKHSA Data Dashboard. The data are available at the national level and are stratified by age group [0-4, 5-14, 15-44, 45-64, 65-79, 80+]. For the purposes of this analysis, age-stratified positivity rates were uniformly averaged to produce a single national weekly series. Part of routine surveillance, influenza and COVID-19 PCR testing data are drawn from laboratories as they report the incidences.

Weekly COVID-19 PCR positivity data were obtained at the regional level, with separate extracts covering the seasons 2022–23, 2023–24, 2024–25, and 2025–26, also from the UKHSA Data Dashboard. Regional data were provided according to older NHS region boundaries and were mapped to the seven current NHS England regions by averaging across constituent sub-regions where necessary (specifically, East and West Midlands were combined into Midlands, and North East and Yorkshire and Humber were combined into North East and Yorkshire).

A further consideration is that SGSS data are subject to retrospective backfilling, meaning that positivity estimates for the most recent days are based on an incomplete set of reported tests and may be revised upward as additional results are processed. While this effect introduces some uncertainty into our positivity estimates, its impact is partially attenuated by our use of positivity rates rather than raw counts, as the ratio of positive to total tests is less sensitive to reporting incompleteness than absolute case counts alone.

#### Hospital Admission Rates

Hospital admissions were included as pathogen-specific indicators of severe disease burden. Daily hospital admission rates for COVID-19, influenza, and RSV were obtained separately for each pathogen at ICB level and subsequently aggregated to the seven NHS England regions. Admission rates were calculated as the number of admissions divided by the corresponding domain population, yielding a per-capita rate.

Admissions are defined as the number of inpatients with a new laboratory-confirmed positive test taken within the past 24 hours. For COVID-19 and influenza, these results come from the Urgent and Emergency Care Daily Situation Report, while RSV results come from SGSS. Estimates of the ICB-level population were calculated via probabilistic mapping using Lower Tier Local Authority (LTLA) and NHS trust boundaries [25]. RSV admission data were available only from approximately 2024 onwards, which is a limitation when interpreting comparisons across pathogens for this study.

#### Data granularity, reporting lag, and regional harmonisation

All analyses were conducted using weekly aggregated time series. Where necessary, surveillance data reported using historical NHS region boundaries were harmonised to the current seven NHS England regions by aggregating constituent sub-regions. Sleep Cycle cough data and hospital admission data were available at Integrated Care Board (ICB) level and were mapped directly to the current NHS regional structure.

Reporting delays are a recognised challenge when evaluating leading indicators in public health surveillance systems [25]. Most of the UKHSA sources considered here have an average reporting delay of approximately 1–2 days, while laboratory-confirmed outcomes may be subject to longer delays. Importantly, these delays are not fixed and may vary over time due to factors such as testing demand, laboratory workload, healthcare utilisation patterns, and seasonal pressures on the healthcare system. We do not adjust any of the surveillance indicators for potential reporting delays, as the actual delay at the time is not available to us, and instead analyse each data source as reported.

### Cross-Correlation Analysis

Let *X_t_* and *Y_t_* denote two time series observed at discrete time points *t* = 1, 2*,…,n*. The cross-correlation function (CCF) is defined at lag *k* as [26]

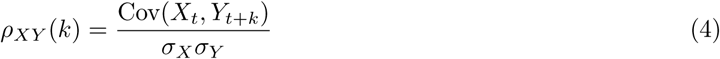

where Cov(*X_t_, Y_t_*_+*k*_) is the covariance between *X_t_* and *Y_t_*_+*k*_, and *σ_X_* and *σ_Y_* are the standard deviations of *X* and *Y* respectively. Note also that *ρ_XY_* (*k*) = *ρ_Y X_* (*−k*) *̸*= *ρ_XY_* (*−k*), to avoid symmetry about *k* = 0.

Expanding the covariance term:

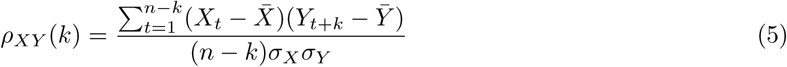

where 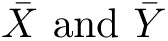 are the sample means of *X_t_* and *Y_t_*. For negative lags (*k <* 0), we compute:

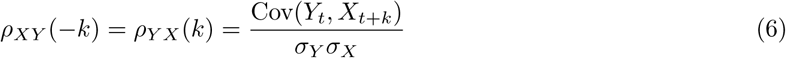

Then,

- When *k* = 0: standard Pearson correlation between *X* and *Y*
- When *k >* 0: correlation between *X_t_* and *Y_t_*_+*k*_ (i.e., *X* leads *Y* by *k* time units)
- When *k <* 0: correlation between *Y_t_* and *X_t−k_* (i.e., *Y* leads *X* by *|k|* time units)

Temporal synchronicity of the time series can be achieved through aligning the earliest start dates and latest end dates of the respective series. Throughout this work, a negative lag *k* indicates that cough activity leads the comparator series by *|k|* time units, such that changes in cough activity are observed *|k|* time units (weeks/days) before corresponding changes in the outcome. Approximate 95% confidence intervals for the prewhitened cross-correlations were calculated as 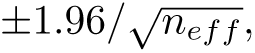 where *n_eff_* is the number of valid paired observations remaining at each lag after prewhitening [27].

### Prewhitening

Cross-correlation analysis can produce misleading lag relationships when the underlying time series exhibit strong autocorrelation or shared seasonal structure. Hendriks et al. [28] demonstrated that apparent lead–lag relationships between influenza-like illness and invasive pneumococcal disease disappeared after accounting for autocorrelation through prewhitening. More generally, correlations between non-stationary time series may arise from shared temporal structure rather than a genuine relationship between the underlying processes [29]. To reduce this risk, we applied prewhitening prior to computing cross-correlations. After successful prewhitening, the residuals should be approximately white noise and therefore any underlying correlation structure that should be accounted for is removed before the cross-correlation.

Prewhitening was performed by removing both seasonal and non-seasonal dependence using seasonal ARIMA(*p, d, q*)(*P, D, Q*)*_s_* models fitted to the cough series *x_t_*:

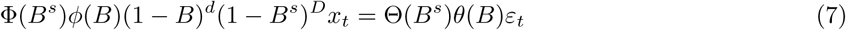

where *B* is the backshift operator, *ϕ*(*B*) and *θ*(*B*) are the autoregressive and moving average polynomials and Φ(*B^s^*) and Θ(*B^s^*) their seasonal counterparts for the seasonal period *s*, and *ε_t_* denotes white-noise residuals. The same filter was then applied to the comparator series *y_t_*:

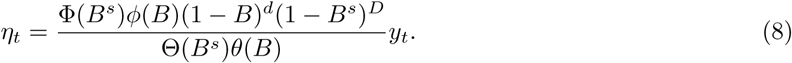

Cross-correlations were subsequently calculated between the filtered series *ε_t_*and *η_t_*:

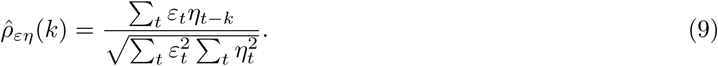

Under this formulation, a significant correlation at lag *k <* 0 indicates that cough activity leads the comparator series by *|k|* weeks, while *k >* 0 indicates the reverse.

ARIMA models were fitted using the auto.arima function from the forecast package in R [30–32]. Model orders were selected automatically using AIC minimisation, with non-stationarity and seasonal terms determined by the fitting procedure. Interpolation of missing values was permitted during model fitting (via na.interp from the forecast package), although this had no practical effect as the cough series contained no missing observations. Figure 7a illustrates successful prewhitening: the autocorrelation structure present in the raw series is largely eliminated after fitting, with autocorrelations at non-zero lags reduced to within the confidence intervals expected under white noise. Figure 7b shows this procedure applied across two related series: the top panels show the cough series being fitted and prewhitened, while the bottom panels show the same ARIMA filter applied to the ARI series, illustrating how a common linear filter is used to remove shared temporal structure prior to cross-correlation. The prewhitened series are flat for the first year shown, reflecting the seasonal differencing term in the fitted model: a full annual cycle of data is required before the model can generate a genuine one-step-ahead residual, so no meaningful signal is produced until this initial conditioning period has elapsed.

**Fig 7.**
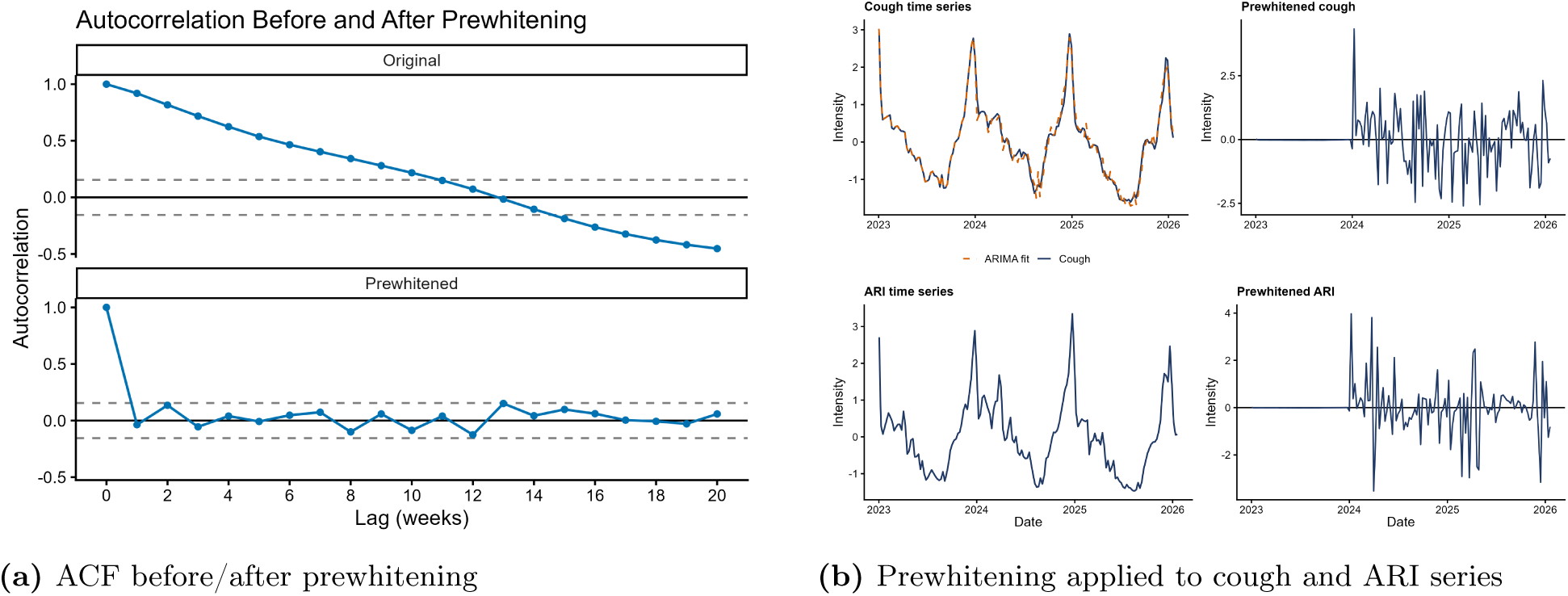
Prewhitening diagnostics for the national weekly average cough rate. Left: autocorrelation function before and after seasonal ARIMA fitting. Right: visualisation of the prewhitening procedure applied to both cough and ARI 111 call series prior to cross-correlation.

### Ethics and privacy

This study used aggregated and anonymised data derived from users of the Sleep Cycle mobile application. No raw audio recordings or personally identifiable information were stored or used during this study. Sleep Cycle shared only aggregated data with UKHSA. UKHSA has legal permission to process confidential patient information for public health purposes under Regulation 3 of the Health Service (Control of Patient Information) Regulations 2002, made under Section 251 of the National Health Service Act 2006. These provisions support the collection, linkage and analysis of identifiable health data for the surveillance, prevention and control of communicable diseases and other threats to public health.

## Data availability statement

The data underpinning this analysis comprise both openly available and protected UKHSA datasets. Openly available data can be obtained from the UKHSA Data Dashboard (https://ukhsa-dashboard.data.gov.uk/) and its associated API. The analysis includes data created through the linkage of multiple health and administrative data sources. Data linkage was undertaken within UKHSA in accordance with relevant legal, ethical, information governance, and data protection requirements. The linked datasets generated and/or analysed during the current study are not publicly available. Protected data used in this study cannot be made publicly available due to legal, ethical, and information governance restrictions. Researchers may apply for access to UKHSA protected data through the UKHSA Data Access Request process, subject to all necessary approvals and disclosure controls (https://www.gov.uk/government/publications/accessing-ukhsa-protected-data/accessing-ukhsa-protected-data). Researchers may request access to the cough dataset via. Requests are evaluated on a case-by-case basis.

## Competing interests

Emil Carlsson and Mikael Kågebäck are employees of Sleep Cycle AB.

## Author contributions

Tommy Irons: Methodology, Software, Formal Analysis, Investigation, Writing – Original Draft, Writing – Review and Editing, Visualisation.

Emil Carlsson: Methodology, Software, Formal Analysis, Investigation, Writing – Original Draft, Writing –

Review and Editing, Visualisation.

Maria Tang: Methodology, Validation, Data Curation.

Jonathon Mellor: Methodology, Validation, Writing – Review and Editing. Conrad Rubin: Data Curation.

Alex Allen: Validation, Writing – Review and Editing. Alex J. Elliot: Validation, Writing – Review and Editing.

Mikael Kågebäck: Supervision, Validation, Writing – Review and Editing.

Josef Packham: Supervision, Project Administration, Funding Acquisition, Writing – Review and Editing.

## Acknowledgments

The authors thank colleagues across UKHSA and partner organisations for their contributions to data collection, curation, linkage, and quality assurance. We acknowledge healthcare providers (including NHS England and NHS 111), laboratories, and surveillance teams whose efforts underpin the data used in this study. A.J.E. is affiliated with the National Institute for Health and Care Research Health Protection Research Unit (NIHR HPRU) in Emergency Preparedness and Response at University of Birmingham, the NIHR HPRU in Gastrointestinal Infections at the University of East Anglia and the NIHR HPRU in Climate Change and Health Security at London School of Hygiene and Tropical Medicine. The views expressed are those of the authors and not necessarily those of NIHR, UKHSA or the Department of Health and Social Care.

## Supporting information

### ARIMA Models

The results of the automatic ARIMA model selection procedure are shown in Table 2. A consistent pattern across regions was that the scaled cough metrics (coughs per user and coughs per hour of sleep) generally required less differencing than total cough counts. This suggests that normalising for variation in user population or sleep duration produced more stable time series, whereas total cough counts retained stronger long-term trends associated with changes in observation volume. Consequently, most models fitted to total cough counts included a first-order differencing term (*d* = 1), while the scaled metrics were typically modelled without differencing (*d* = 0). All selected models included autoregressive components, indicating temporal dependence in cough activity across weeks. Moving-average terms were included where supported by the data through the automatic model selection procedure.

**Table 2.** Prewhitening ARIMA models fitted to each cough metric series. Models are reported in full seasonal form (*p, d, q*)(*P, D, Q*)[*s*].

| Geography | Frequency | Group | Total | Hourly Avg | Per User |
| --- | --- | --- | --- | --- | --- |
| National | Weekly | — | (0,1,1)(0,1,0)[52] | (2,1,1)(1,1,0)[52] | (0,1,3)(1,1,0)[52] |
| Regional | Weekly | East of England | (1,1,1)(0,1,0)[52] | (0,1,0)(1,1,0)[52] | (0,1,3)(1,1,0)[52] |
| Regional | Weekly | London | (0,1,1)(0,1,0)[52] | (0,1,1)(0,1,1)[52] | (0,1,1)(0,1,1)[52] |
| Regional | Weekly | Midlands | (0,1,1)(0,1,0)[52] | (0,1,2)(0,1,1)[52] | (3,1,0)(0,1,1)[52] |
| Regional | Weekly | North East and Yorkshire | (0,1,1)(0,1,0)[52] | (0,1,3)(0,1,1)[52] | (0,1,3)(0,1,1)[52] |
| Regional | Weekly | North West | (0,1,1)(0,1,0)[52] | (0,1,0)(1,1,0)[52] | (2,0,0)(1,1,0)[52] |
| Regional | Weekly | South East | (1,1,1)(0,1,0)[52] | (2,1,1)(0,1,1)[52] | (0,1,3)(0,1,1)[52] |
| Regional | Weekly | South West | (2,1,2)(0,1,0)[52] | (1,1,1)(1,1,0)[52] | (1,1,1)(1,1,0)[52] |

## Notes

### Competing Interest Statement

Emil Carlsson and Mikael Kageback are employees of Sleep Cycle AB.

### Author Declarations

UKHSA have an exemption under regulation 3 of Sect. 251 of the National Health Service Act (2006) to allow identifiable patient information to be processed to diagnose, control, prevent, or recognise trends in, communicable diseases and other risks to public health. The data are non-identifiable aggregate patient data. I confirm that all necessary patient/participant consent has been obtained and the appropriate institutional forms have been archived, and that any patient/participant/sample identifiers included were not known to anyone (e.g., hospital staff, patients or participants themselves) outside the research group so cannot be used to identify individuals. For this research, Sleep Cycle shares only aggregated data protected using differential privacy. As such, no personal data or information that could reasonably identify individual users are shared between the parties. The data originate from Sleep Cycle users who have accepted the app's Terms & Conditions, Privacy Notice, and any applicable user consents required for the processing of their data.

